# Selecting donors for faecal microbiota transplantation in ulcerative colitis

**DOI:** 10.1101/2020.03.25.20043182

**Authors:** Marcel A. de Leeuw, Manuel X. Duval

## Abstract

**Background:** Inflammatory bowel disease (IBD) is a set of conditions characterized by non-infectious chronic inflammation of the gastrointestinal tract. These primarily include Crohn’s disease (CD), ulcerative colitis (UC) and indeterminate colitis. Fecal microbiota transplantation (FMT) has proven to be an effective treatment for some patients with active UC. There is currently no procedure allowing to predict the patients’ response and to select the most adequate donor(s).

**Aim:** Investigate microbiome characteristics in association with responder/non-responder status and develop selection criteria for donor samples to be used for UC FMT.

**Methods:** Available UC longitudinal FMT microbiome data sets were in part combined and reanalyzed, with focus on species level changes in the microbiota, using state-of-the-art 16S analysis routines.

**Results:** We predicted antibiotic resistance to be higher in non-responders (p=0.0064). Microbiomes of UC FMT responder donors have higher phylogenetic diversity (p=0.0026) and a higher proportion of facultative anaerobes (p=3.3E-5) as compared to non-responder donors. We predicted succinate and histamine to be increased in non-responder donors and non-responders, respectively. Sialic acid catabolism was also predicted to be increased in non-responder donors. Tryptamine and indole-3-acetaldehyde were predicted to be increased in responder donors.

**Conclusions:** Our findings contribute to the establishment of selection criteria for UC FMT donor samples and composition guidelines for future synthetic microbial communities. Our results suggest that oxidative stress resistant facultative anaerobes are important for the establishment of an anaerobic environment and a successful UC FMT therapy. Several metabolites can be tested for additional optimization or prioritization of stool bank samples for UC FMT. Our results question the usefulness of antibiotics based preparation of the gut, prior to FMT.

## Introduction

The incidence and prevalence of inflammatory bowel disease (IBD) are increasing worldwide. It can affect people of all ages, including children and geriatric populations, and can impact all aspects of life. The Global number of cases of IBD reached 6.8 million in 2017, with the highest age-standardized prevalence rates in the U.S.A. and the U.K. [1]. IBD is characterized by non-infectious chronic inflammation of the gastrointestinal tract, and primarily includes Crohn’s disease (CD), ulcerative colitis (UC) and indeterminate colitis [2]. Whereas CD can affect any part of the gastroesophageal tract from the mouth to the anus, UC is confined to the large bowel. Diagnosis of IBD relies on a combination of medical history, physical examination, laboratory testing, and endoscopy with biopsy [3]. Treatment goals for IBD are to minimize symptoms, improve quality of life, and minimize progression and complications of the disease. Most treatments for UC target the immune system through mediators of the inflammatory cascade [4], but a substantial proportion of UC patients continue to have inadequate disease control.

Over 230 genetic risk loci have been associated with IBD. The association of the nucleotide-binding oligomerization domain containing protein 2 (NOD2), an intra-cellular innate immune sensor of bacteria, remains the strongest association to date [5]. The microbiota of UC patients are characterized by reduced complexity with depletion of commensal *Firmicutes* and *Bacteroidetes* [6–11], which is reflected in IgG antibody composition [12]. Mucosal concentration of bacteria is markedly increased in IBD [13], which is further accentuated in UC [14].

Fecal microbiota transplantation (FMT) is a therapeutic procedure aimed at replacing dysbiotic microbiota with physiological/commensal species by the administration of filtered fecal material from a healthy donor into the intestinal tract of a patient [15]. A number of studies, including randomized controlled trials [16–20], suggest that FMT is inducing remission in active UC, for approximately one out of three patients [21]. It is as yet unclear which donor and recipient related factors contribute to a successful clinical response, although in one study, donor stool diversity reached significance [17]. Current consensus guidelines focus on reducing the risk of potentially transmittable disease [22]. Overall, FMT for UC needs more specification and more supporting evidence of long term efficacy and safety before being recommended for clinical practice [21, 23].

We re-used a series of relevant longitudinal microbiome studies, available in the form of raw 16S data, to address elementary gut microbiome composition pertaining to clinical endpoints. Where possible, we combined datasets to increase statistical power. We initiated our study by describing the dysbiosis of UC in the broader context of immune mediated inflammatory diseases (IMIDs). Next we investigated compositional and functional differences between responder- and non-responder patient and donor gut microbiomes.

## Materials & Methods

The materials summarized in Table 1 have been made available as part of clinical trials. All data sets are longitudinal, with data points covering pre-FMT assessment and post-FMT follow-up. Only one available data set, SRP254202, was eliminated entirely because of a high amount of non-specific priming. Biopsy samples from data set ERP116682 were eliminated because we detected high amounts of species reported as reagent contaminants [24]. Patient stool samples from four studies with available responder/non-responder status were combined to increase statistical power. Screening and post-FMT samples from ERP013257, SRP135559, ERP116682 and SRP102742 comprised 46 responders and non-responders to FMT and 159 samples. Partial responders from ERP013257 were discarded to homogenize endpoints. Studies SRP135559 and SRP102742 comprised children, whereas ERP013257 was from an adult population. Studies SRP135559 and SRP102742 used pediatric ulcerative colitis activity index (PUCAI) scores to assess response, whereas ERP013257 and ERP116682 used Mayo scores to do so.

**Table 1:** Longitudinal UC FMT data sets used in the study. N: number of patients, n: number of samples, 16S: variable regions covered, *:unpublished.

| BioProject | SRA | N | n | type | 16S | publ. |
| --- | --- | --- | --- | --- | --- | --- |
| PRJEB11841 | ERP013257 | 27 | 172 | stool | V4 | [17] |
| PRJEB26474 | ERP108465 | 81 | 158 | biopsy | V1-V2 | [19] |
| PRJEB26472 | ERP108463 | 81 | 309 | stool | V1-V2 | [19] |
| PRJNA515212 | SRP180003 | 7 | 42 | stool | V3-V4 | [25] |
| PRJEB33851 | ERP116682 | 28 | 98 | stool | V4 | * |
| PRJNA475599 | SRP198502 | 12 | 235 | stool | V4 | [26] |
| PRJNA380944 | SRP102742 | 21 | 109 | stool | V4 | [27] |
| PRJNA438164 | SRP135559 | 7 | 21 | stool | V4 | [28] |
| PRJNA388210 | SRP108284 | 20 | 78 | stool | V4 | * |
| total |  | 203 | 1,222 |  |  |  |

For the assessment of donor microbiome characteristics in association with response, we disposed of but one study, ERP013257, comprising 12 donors and 51 donor samples. We used the partial response versus remission distinction provided by this study [17].

For the purpose of profiling ulcerative colitis and Crohn’s disease microbiomes, we also used the published data set listed in supplemental Table S1.

### Data analysis

Amplicon Sequence Variants (ASVs) were generated with the R Bioconductor package dada2 [29], version 1.12.1 with recommended parameters, involving quality trimming, discarding of sequences with N’s, assembly of forward and reverse sequences and chimera removal, as described previously [30]. Further analysis involved multiple alignment with mafft, version 6.603b [31] and approximately-maximum-likelihood phylogenetic tree generation with FastTreeMP, version 2.1.11 [32], both with default settings.

Taxonomic classification of ASVs was performed by an in-house Python and R program using random forest based supervised learning on the Ribosomal Database Project (RDP) release 11.5. Phenotypic content of microbiomes was inferred using species-level taxonomic assignments only and an in-house phenotypic database which has been initiated with information from BacDive [33] and the IJSEM journal [34] and has been hand-curated since. Resulting classifications and microbiome phenotypes are available from the Github repository https://github.com/GeneCreek/UC-manuscript in the form of Phyloseq R data objects.

UniFrac distances were computed using the R Bioconductor package phyloseq, version 1.28.0 [35] on raw ASVs. Further analysis used counts and relative abundances summarized at the species level, using the taxonomic classifications.

Detection of responder and non-responder associated taxa from a prevalence perspective was performed with `χ` ^2^ testing, with Monte Carlo simulation based computation of p-values [36].

Detection of responder and non-responder associated taxa from a relative abundance perspective was performed with the R package DESeq2, using negative binomial GLM fitting as described previously [30], requiring pAdj < 0.001.

Relative gene abundance prediction was performed using in-house R code, correcting for 16S copy numbers using the rrnDB version 5.4 [37] and using UniProt [38] provided per species non-redundant proteome counts and per species gene counts. In brief, UniProt queries were constructed to target key genes in pathways of interest. Resulting gene counts per species were normalized w.r.t. the number of available non-redundant proteomes for each species and multiplied by the rrnDB normalized relative abundance of species in the experimental data. Tukey honest significant difference testing was applied to the resulting gene abundance estimates to investigate difference between sample groups. This approach is similar to the Picrust algorithm [39] but is not limited to clusters of orthologous genes (COGs).

Downstream analysis scripts covering the figures presented in the manuscript are available from https://github.com/GeneCreek/UC-manuscript.

## Results

### Dysbiosis in IMIDs

Data set SRP183770 contains biological replicates of stool samples collected from patients with Crohn’s disease (CD; N = 20), ulcerative colitis (UC; N = 19), multiple sclerosis (MS; N = 19), and rheumatoid arthritis (RA; N = 21) versus healthy controls (HC; N = 23) [40]. We initiated our study by describing the distribution of two fecal microbiome phenotypes, the species diversity with the Shannon diversity index and the strict anaerobes proportion, in UC samples within the context of these other IMIDs, Fig. 1. The strict anaerobes proportion is estimated as the proportion of obligate anaerobes among anaerobes. The ranking of the IMID’s diseases is the same according to these two phenotypes, with notably rheumatoid arthritis positioned between ulcerative colitis and Crohn’s disease.

**Figure 1:**
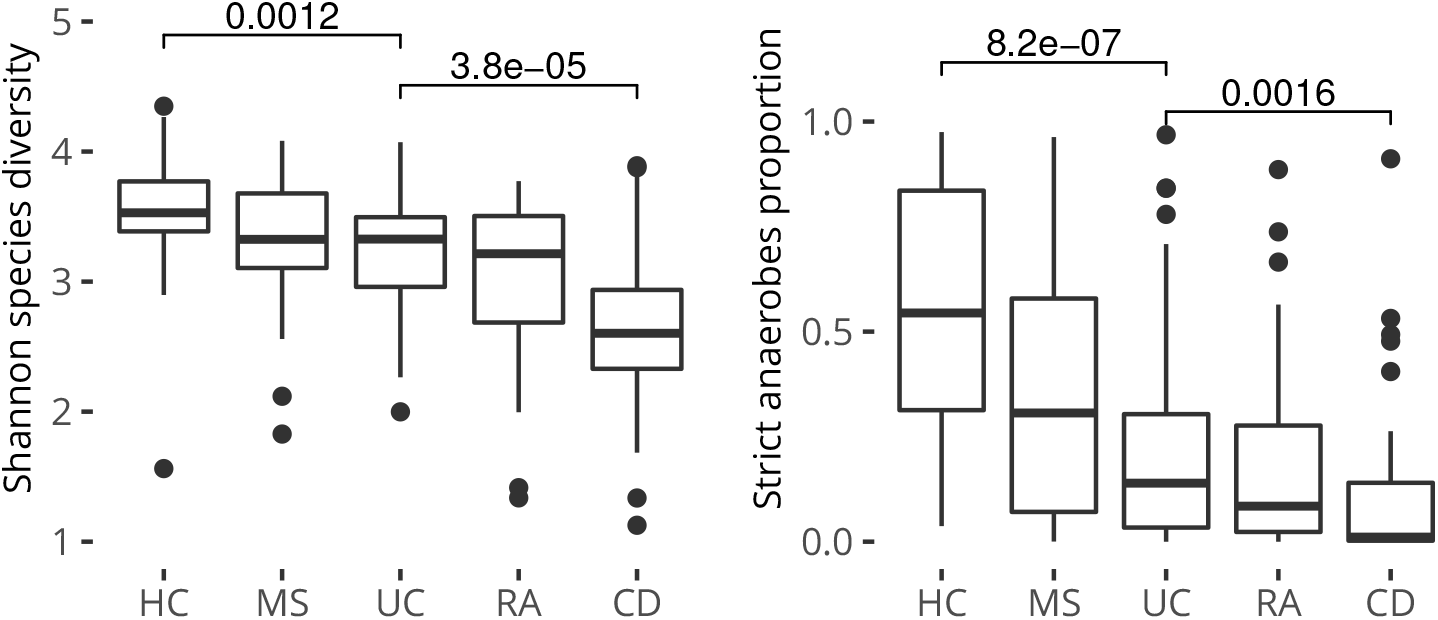
Microbiome phenotypes in IMIDs. Data set SRP183770, 79 subjects, 201 samples. Numbers reflect Wilcoxon signed rank test continuity corrected p-values. HC: healthy controls, MS: multiple sclerosis, UC: ulcerative colitis, RA: rheumatoid arthritis, CD: Crohn’s disease.

Several publications have reported dysbiosis in UC can be reduced through FMT. This principle is illustrated using data set SRP108284 (20 patients) and Chao species richness in supplemental Fig. S1. The improvement reaches statistical significance in the samples collected in the second week after a single FMT, but washes out subsequently in the fourth week.

Just like species diversity, the strict anaerobe proportion can be modified through FMT, Fig. S2. The two donors used in study SRP198502 (7 patients) were selected based on fecal butyrate concentration [26] and had a high proportion of strict anaerobes.

### Responder and non-responder patients

Combining the UC patient gut microbiomes from all four data sets for which responder/non-responder status was available in study metadata, we tested several microbiome phenotypes in relation to the responder status, two of which reached significance, Fig. 2. Responders have a lower predicted average biological safety level (BSL) and a predicted more gram positive gut microbiome.

**Figure 2:**
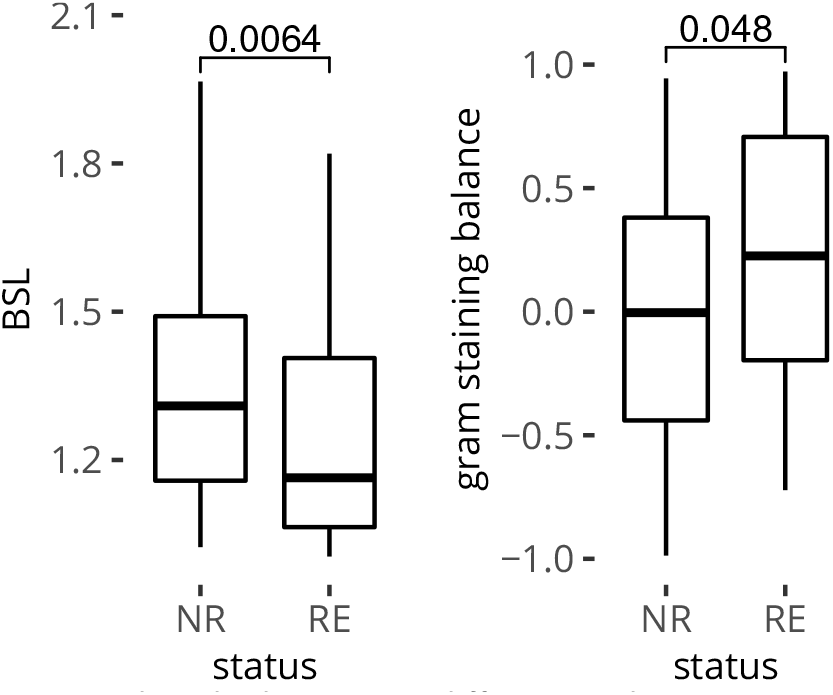
Predicted phenotypic differences between responder and non-responder microbiomes. Combined data sets ERP013257, SRP135559, ERP116682 and SRP102742, 46 patients, 159 samples. BSL: predicted average biological safety level of the microbiome. NR: non-responders, RE: responders. Numbers reflect Wilcoxon signed rank test continuity corrected p-values.

In data set ERP013257 [17] the predicted average BSL of the microbiome is significantly different between non-responders, partial responders and responders, Fig. S3. In the experience, an antibiotics course was used prior to FMT. In non-responders and partial responders, there was a spike in BSL after the 10-day antibiotics course, whereas in responders, this phenomenon was not seen. Subsequently, after FMT, the BSL remained lower in responders, Fig. 3.

**Figure 3:**
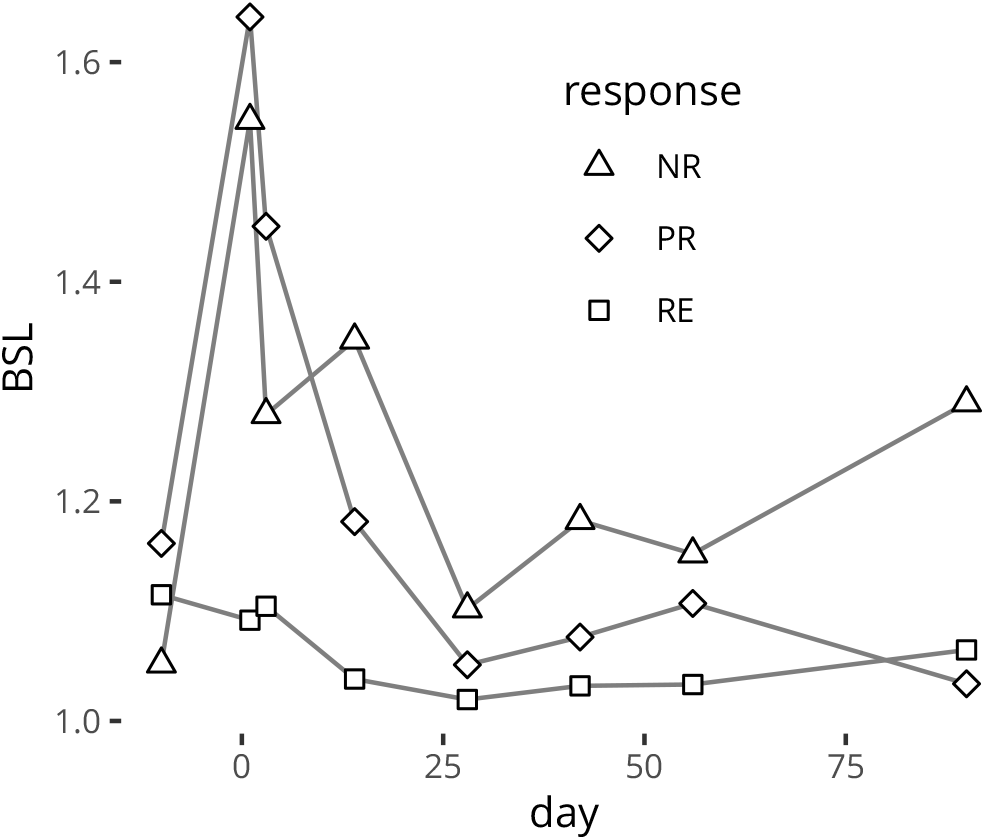
BSL trajectory across antibiotics course, FMT and follow-up. Data set ERP013257, 13 patients. Data points reflect the median predicted biological safety levels per response group. NR: non-responders, PR: partial responders, RE: responders. Antibiotics course: days −10 to 0, FMT: day 1.

We also computed both prevalence and relative abundance differences between non-responders and responders using the four responder/non-responder data sets combined, Table S2 respectively S3 and S4. We found *Allisonella histaminiformans* to be the most significantly prevalent species in non-responders.

### Responder and non-responder donors

Data set ERP013257 comprises 51 samples from 12 donors which are associated with either non-response, partial response or response to UC treatment with FMT [17]. Multi-dimensional scaling ordination separates responders- and non-responder donors, with partial responder donors in an intermediate position, Fig. S4. We tested several microbiome phenotypes, in relation to the responder donor status. Phylogenetic diversity and predicted average oxygen tolerance reached significance, Fig. 4.

**Figure 4:**
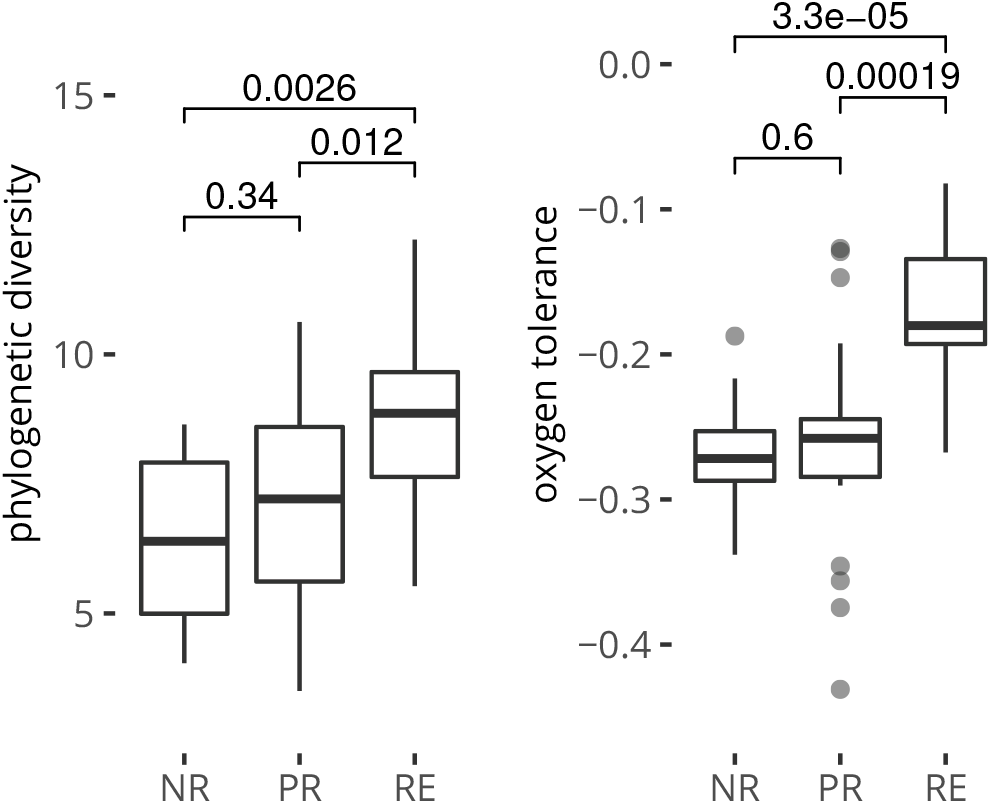
Donor microbiome phenotypes, data set ERP013257. 51 samples from 12 donors. Numbers reflect Wilcoxon signed rank test continuity corrected p-values. Donors are associated with NR: non-responders, PR: partial responders, RE: responders.

We also computed prevalence and relative abundance differences between non-responder and responder donors using data set ERP013257, Table S5 respectively S6. We found very little overlap in composition between donor- and patient responders or non-responders.

### Functional diferences between responder and non-responder microbiomes

The following metabolites in relation to UC were the subject of gene queries to the UniProtKB section of UniProt: succinate, glutathione, tryptophan and sialic acid. Queries provided in Table S7 gave rise to significant differences between responder and non-responder microbiomes or between responder and non-responder donor microbiomes as appreciated by Tukey honest significant difference testing, Table 2.

**Table 2:**
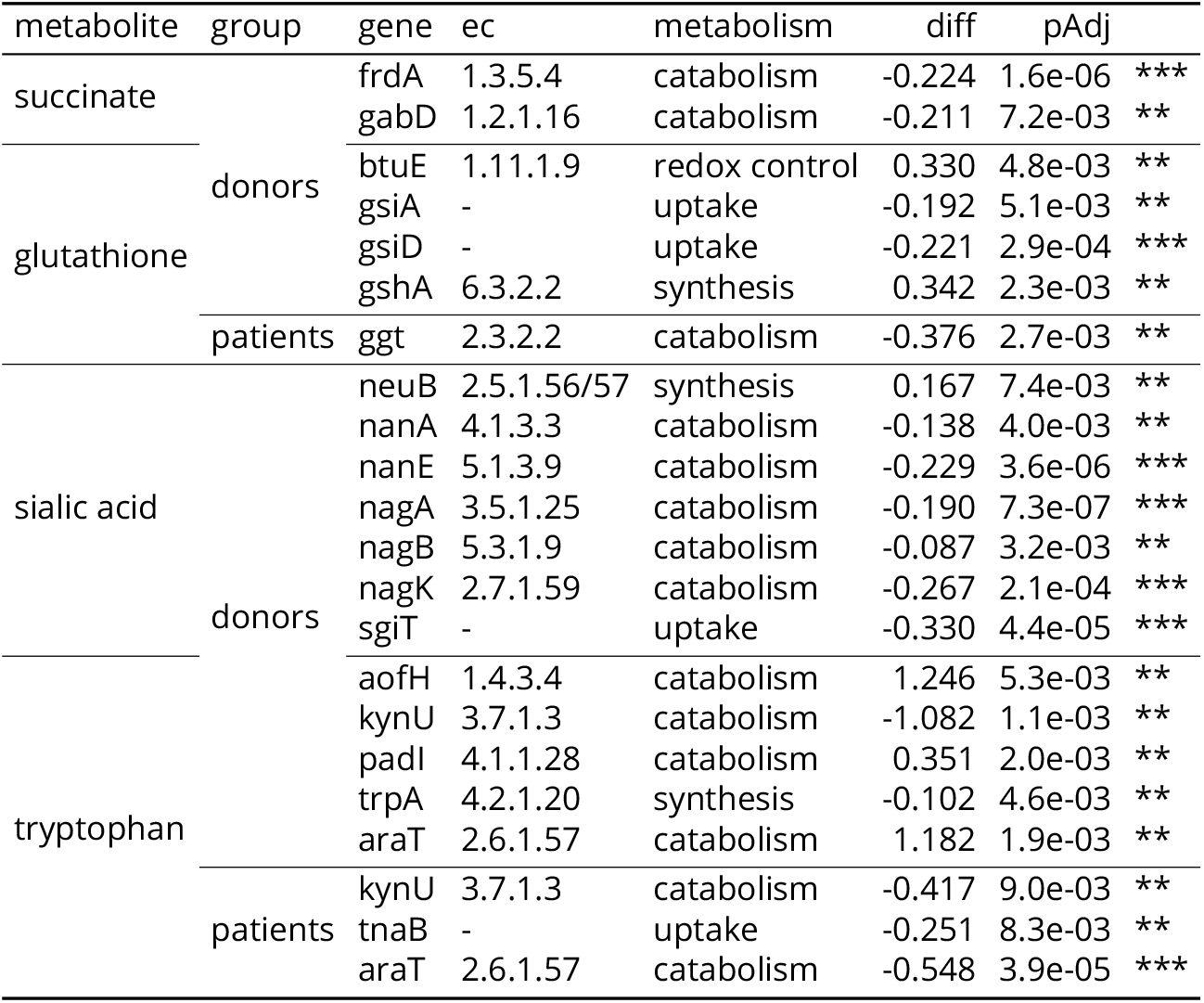
Predicted functional differences. Tukey honest significant difference test results on univariate linear models of predicted relative gene abundance. Comparison: responders − non-responders. ec: enzyme commission number. aofH: monoamine oxidase, araT: aromatic amino acid transaminase, btuE: glutathione peroxidase, frdA: fumarate reductase, gabD: succinate-semialdehyde dehydrogenase, ggt: glutathione hydrolase, gshA: glutathione biosynthesis bifunctional, gsiA: glutathione import ATP-binding, gsiD: glutathione permease, kynU: kynureninase, nagA: N-acetylglucosamine-6-phosphate deacetylase, nagB: glucosamine-6-phosphate isomerase, nagK: N-acetylglucosamine kinase, nanA: sialic acid aldolase, nanE: acylglucosamine P epimerase, neuB: NeuAc synthase, padI: aromatic acid decarboxylase, sgiT: putative sodium sialic acid symporter, tnaB: tryptophan permease, trpA: tryptophan synthase alpha

Succinate is a metabolic intermediate of the tricarboxylic acid (TCA) cycle. Succinate is also produced in large amounts during bacterial fermentation of dietary fiber. Fecal succinate concentrations are approximately three to four-fold higher in IBD patients compared to controls [41]. When projecting relative gene abundance, we found succinate producing fumarate reductase and succinate-semialdehyde dehydrogenase associated with non-responder donors.

Glutathione (GSH) is one of the most potent natural antioxidants and found mostly in Gram-negative bacteria [42]. GSH metabolism involves several genes including catabolism (amino-acid uptake), synthesis and use for redox control. Projecting relative gene abundance, we found several genes were predicted to be differentially abundant between responder- and non-responder donors. Synthesis and use for redox control were associated with responder donors and uptake was associated with non-responder donors. When comparing predicted relative gene abundance in patient microbiomes, one additional catabolic gene, ggt, was found associated with non-responders.

Tryptophan metabolism has been found associated with IBD activity, with possible implication of gut microbiota [43]. We found three tryptophan metabolism genes, one uptake and two catabolic genes predicted to be increased in non-responder patients, Table 2. We found the first catabolic gene, kynU in association with non-responder donors, but paradoxically the second catabolic gene, araT, in association with responder donors.

Sialic acid is an abundant and the most terminally expressed carbohydrate on mucus glycans. Bacterial metabolism of sialic acid comprises scavenging, catabolism, synthesis and decoration [44]. Projecting relative gene abundance, we found several genes are predicted to be differentially abundant between responder- and non-responder donors. Catabolism was found associated with non-responder donors and synthesis with responder donors.

## Discussion

### Limitations

The majority of the data sets used in this study are from 16S variable region V4 sequencing. A substantial number of species cannot be resolved using this amplicon. A better alternative is the combined V3-V4 amplicon which can be sequenced using paired-end reads.

### Comparison with previous findings

We found the Shannon species diversity to be the most discriminant diversity or richness criterium to separate the IMIDs as reported previously [40]. The ranking we found is also identical. The equally discriminant strict anaerobe proportion differences between IMIDs we found were not investigated in that publication.

In the four studies we combined to investigate gut microbiome differences between UC FMT responders and non-responders, the biological safety level and the gram staining balance have not been investigated. Without combination of the studies, the gram staining balance difference we reported discriminant did not reach significance.

We found UC FMT responder donor samples had higher phylogenetic diversity as compared to non-responder donors. This is in line with findings from the original publication of data set ERP013257 [17]. Microbiome oxygen tolerance was not investigated in that publication.

In none of the original publications, prediction of functional differences between UC FMT responders and nonresponders or responder donors and non-responder donors has been performed.

### Oxidative stress in UC

Oxidative stress has been proposed as a mechanism underlying the pathophysiology of CD [45] and has long been known as a pathogenic factor of UC [46]. If radical oxygen species (ROS) are released in the lumen, we may expect modulation of the microbiome composition. Indeed, we observe a decreased proportion of strict anaerobes in the microbiome composition of UC and CD samples, accompanied by loss of species diversity. The loss of obligate anaerobes in IBD has been reported previously [47]. Thus we postulate oxidative stress in IMIDs drives the decrease in obligate anaerobes, which results in the loss of microbial diversity and loss of function such as butyrate production.

Notwithstanding the above, we found responder donor samples had higher predicted oxygen tolerance as compared to non-responder donors. Thus rather than directly supplying missing strict anaerobes, provision of facultative anaerobes capable of restoring an anaerobic environment seems important for successful UC FMT.

### Antibiotics based gut preparation

We found a remarkable association between predicted microbiome BSL, i.e. antibiotic resistance and non-responder/responder status, with responder microbiomes having a lower predicted BSL. This finding highlights the controversial role of broad-spectrum antibiotics based preparation of the gut prior to FMT [48], if applied. The positive selection of resistant species seems to have a detrimental effect on FMT success. It appears from our findings that antibiotics based cleansing in the specific context of UC in addition to conventional bowel cleaning solution used for colonoscopy preparation needs to be questioned.

### Metabolites and UC

Increased intestinal succinate levels have been reported in the well-known Dextran Sulfate Sodium (DSS) induced mouse colitis model and proposed as a causative agent [49]. In humans, fecal succinate concentrations are approximately three to four-fold higher in IBD patients compared to controls [41]. We found predicted relative gene abundance of two succinate producing enzymes increased in non-responder donors.

Mucosal histamine has been reported to reflect the degree of colonic inflammation in Crohn’s disease and ulcerative colitis [50]. Furthermore, the His645Asp polymorphism of the histamine metabolizing enzyme ABP1 is related to severity of ulcerative colitis [51]. We found *Allisonella histaminiformans* to be the most differentially prevalent species in non-responders. We also found histamine forming histidine decarboxylase (hdc, ec: 4.1.1.22) carriers such as *Clostridium perfringens, Lactobacillus reuteri, Proteus vulgaris* and *Morganella morganii* among non-responder associated species. Reportedly, *Enterobacter cloaceae* and *Proteus mirabilis* are also histamine producers [52]. We found both associated with non-responders too. Histidine decarboxylase did, however, not reach significance in our predicted relative gene abundance test.

Decreased serum levels of tryptophan are correlated with disease activity in IBD [43]. Tryptophan synthesis is a virulence factor of bacterial pathogens, allowing for survival in macrophages. We found tryptophan synthase A to be predicted increased in non-responder donors. It is more intricate to interpret predictions regarding the metabolite indole-3-pyruvic acid (IPA), which we find associated with non-responders but also with responder donors. The latter is coherent with the fact that IPA is diminished in circulating serum from human subjects with active colitis [53]. One possible explanation lies in species specific substrate preferences of aromatic amino acid transaminase, which is potentially also active on tyrosine and phenylalanine. Lastly, we found two catabolic genes associated with responder donors, padI and aofH, respectively and successively producing tryptamine and indole-3-acetaldehyde (IAAld), both aryl hydrocarbon receptor (AhR) ligands contributing to AhR-dependent IL-22 transcription [54]. AhR ligands have also been shown to regulate the IL-10 receptor [53]. Thus these products contribute to intestinal homeostasis.

### Mucosal barrier function

Sialic acid is an abundant and the most terminally expressed carbohydrate on mucus glycans. Removal of sialic acid is the first overall step in microbial infiltration of the mucus layer. Exosialidases have been shown to increase availability of nutrients for *Escherichia coli* expansion in the DSS induced mouse colitis model [55]. We found predicted sialic acid uptake and catabolism but not scavenging in strong association with non-responder donors. The various *Bacteroides* species, *Akkermansia muciniphyla* and *Feacilibacterium prausnitzii* carrying the scavenger gene nanH were found equally present in responder- and non-responder patients and donors.

### Prospects

Our findings can be used to rationalize selection of donor samples from stool banks. Transplantation of facultative anaerobes can be enhanced with selection of donor samples through 16S sequencing as demonstrated. From a metabolic standpoint, low succinate and histamine levels should be preferred, whereas higher tryptamine and IAAld levels would be a plus. Sialic acid catabolism could be addressed through qPCR of key catabolic enzymes encoding genes such as nanH and nanA, of which lower levels would reduce over-colonization of the mucus barrier. Our results suggest that in the specific context of UC FMT, broad-spectrum antibiotics based preparation of the gut needs to be compared in a randomized clinical trial against solution based cleansing only.

### Conclusions

In conclusion, we found the microbiomes of UC and CD patients to have paucity of obligate anaerobes, which could be related to oxidative stress. Microbiomes of UC FMT responders and responder donors have higher phylogenetic diversity and a higher proportion of facultative anaerobes as compared to non-responders and non-responder donors. We found several predicted metabolites associated with either non-responder status or responder status of patients and donors. We also found predicted sialic acid catabolism to be significantly associated with non-responder donor status. These findings should contribute to the establishment of selection criteria for UC FMT donor samples and composition guidelines for future synthetic microbial communities.

## Data Availability

All data and scripts required to reproduce the analysis are available from the following GitHub archive https://github.com/GeneCreek/UC-manuscript

## Declarations

## Acknowledgements

The authors received no financial support for this study. The authors acknowledge the contributions to the Short Read Archive made by the respective institutions and acknowledge scientific journals for enforcing this practice.

## Authors’ contributions

Study design, data collection, data analysis and writing of the manuscript (ML); data analysis and writing of the manuscript (MD).

## Bibliography

[1] GBD 2017 IBD Collaborators. The global, regional, and national burden of inflammatory bowel disease in 195 countries and territories, 1990-2017: a systematic analysis for the Global Burden of Disease Study 2017. The Lancet Gastroenterology & Hepatology, 5(1):17–30, January 2020. doi:10.1016/S2468-1253(19)30333-4.

[2] Barbara A Hendrickson, Ranjana Gokhale, and Judy H Cho. Clinical aspects and pathophysiology of inflammatory bowel disease. Clinical Microbiology Reviews, 15(1):79–94, January 2002. doi:10.1128/cmr.15.1.79-94.2002.

[3] Tomoko Sairenji, Kimberly L Collins, and David V Evans. An Update on Inflammatory Bowel Disease. Primary care, 44(4):673–692, December 2017. doi:10.1016/j.pop.2017.07.010.

[4] Grainne Holleran, Loris Lopetuso, Valentina Petito, Cristina Graziani, Gianluca Ianiro, Deirdre McNamara, Antonio Gasbarrini, and Franco Scaldaferri. The Innate and Adaptive Immune System as Targets for Biologic Therapies in Inflammatory Bowel Disease. International journal of molecular sciences, 18(10), September 2017. doi:10.3390/ijms18102020.

[5] Ashleigh Goethel, Kenneth Croitoru, and Dana J Philpott. The interplay between microbes and the immune response in inflammatory bowel disease. The Journal of physiology, 596(17):3869–3882, September 2018. doi:10.1113/JP275396.

[6] Daniel N Frank, Allison L St Amand, Robert A Feldman, Edgar C Boedeker, Noam Harpaz, and Norman R Pace. Molecular-phylogenetic characterization of microbial community imbalances in human inflammatory bowel diseases. PNAS, 104(34): 13780–13785, August 2007. doi:10.1073/pnas.0706625104.

[7] Shadi Sepehri, Roman Kotlowski, Charles N Bernstein, and Denis O Krause. Microbial diversity of inflamed and noninflamed gut biopsy tissues in inflammatory bowel disease. Inflammatory Bowel Diseases, 6(6):675–683, June 2007. doi:10.1002/ibd.20101.

[8] Samah O Noor, Karyn Ridgway, Louise Scovell, E Katherine Kemsley, Elizabeth K Lund, Crawford Jamieson, Ian T Johnson, and Arjan Narbad. Ulcerative colitis and irritable bowel patients exhibit distinct abnormalities of the gut microbiota. BMC gastroenterology, 10(1):134–9, November 2010. doi:10.1186/1471-230X-10-13.

[9] Mirjana Rajilić-Stojanović, Fergus Shanahan, Francisco Guarner, and Willem M de Vos. Phylogenetic analysis of dysbiosis in ulcerative colitis during remission. Inflammatory Bowel Diseases, 19(3):481–488, March 2013. doi:10.1097/MIB.0b013e31827fec6d.

[10] Fei Sjöberg, Cecilia Barkman, Intawat Nookaew, Sofia Östman, Ingegerd Adlerberth, Robert Saalman, and Agnes E Wold. Low-complexity microbiota in the duodenum of children with newly diagnosed ulcerative colitis. PLoS ONE, 12(10), 2017. doi:10.1371/journal.pone.0186178.

[11] Hengameh Chloé Mirsepasi-Lauridsen, Katleen Vrankx, Jørgen Engberg, Alice Friis-Møller, Jørn Brynskov, Inge Nordgaard-Lassen, Andreas Munk Petersen, and Karen Angeliki Krogfelt. Disease-Specific Enteric Microbiome Dysbiosis in Inflammatory Bowel Disease. Frontiers in medicine, 5, 2018. doi:10.3389/fmed.2018.00304.

[12] E Furrie, S Macfarlane, J H Cummings, and G T Macfarlane. Systemic antibodies towards mucosal bacteria in ulcerative colitis and Crohn’s disease differentially activate the innate immune response. BMJ Gut, 53(1):91––98, January 2004. doi:10.1136/gut.53.1.91.

[13] Alexander Swidsinski, Axel Ladhoff, Annelie Pernthaler, Sonja Swidsinski, Vera Loening-Baucke, Marianne Ortner, Jutta Weber, Uwe Hoffmann, Stefan Schreiber, Manfred Dietel, and Herbert Lochs. Mucosal flora in inflammatory bowel disease. Gastroenterology, 1(1):44–54, January 2002. doi:10.1053/gast.2002.30294.

[14] Rodrigo Bibiloni, Marco Mangold, Karen L Madsen, Richard N Fedorak, and Gerald W Tannock. The bacteriology of biopsies differs between newly diagnosed, untreated, Crohn’s disease and ulcerative colitis patients. Journal of medical microbiology, 55(Pt 8):1141–1149, August 2006.

[15] Faming Zhang, Bota Cui, Xingxiang He, Yuqiang Nie, Kaichun Wu, Daiming Fan, and FMT-standardization Study Group. Microbiota transplantation: concept, methodology and strategy for its modernization. Protein & cell, 9(5):462–473, May 2018. doi:10.1007/s13238-018-0541-8.

[16] Paul Moayyedi, Michael G Surette, Peter T Kim, Josie Libertucci, Melanie Wolfe, Catherine Onischi, David Armstrong, John K Marshall, Zain Kassam, Walter Reinisch, and Christine H Lee. Fecal Microbiota Transplantation Induces Remission in Patients With Active Ulcerative Colitis in a Randomized Controlled Trial. Gastroenterology, 149(1):102– 109.e6, July 2015. doi:10.1053/j.gastro.2015.04.001.

[17] P Kump, P Wurm, H P Gröchenig, H Wenzl, W Petritsch, B Halwachs, M Wagner, V Stadlbauer, A Eherer, K M Hoffmann, A Deutschmann, G Reicht, L Reiter, P Slawitsch, G Gorkiewicz, and C Högenauer. The taxonomic composition of the donor intestinal microbiota is a major factor influencing the efficacy of faecal microbiota transplantation in therapy refractory ulcerative colitis. Alimentary Pharmacology & Therapeutics, 47(1):67–77, October 2017. doi:10.1111/apt.14387.

[18] Sudarshan Paramsothy, Michael A Kamm, Nadeem O Kaakoush, Alissa J Walsh, Johan van den Bogaerde, Douglas Samuel, Rupert W L Leong, Susan Connor, Watson Ng, Ramesh Paramsothy, Wei Xuan, Enmoore Lin, Hazel M Mitchell, and Thomas J Borody. Multidonor intensive faecal microbiota transplantation for active ulcerative colitis: a randomised placebo-controlled trial. Lancet (London, England), 10075(10075):1218–1228, March 2017. doi:10.1016/S0140-6736(17)30182-4.

[19] Sudarshan Paramsothy, Shaun Nielsen, Michael A Kamm, Nandan P Deshpande, Jeremiah J Faith, Jose C Clemente, Ramesh Paramsothy, Alissa J Walsh, Johan van den Bogaerde, Douglas Samuel, Rupert W L Leong, Susan Connor, Watson Ng, Enmoore Lin, Thomas J Borody, Marc R Wilkins, Jean-Frederic Colombel, Hazel M Mitchell, and Nadeem O Kaakoush. Specific Bacteria and Metabolites Associated With Response to Fecal Microbiota Transplantation in Patients With Ulcerative Colitis. Gastroenterology, 156(5):1440–1454.e2, April 2019. doi:10.1053/j.gastro.2018.12.001.

[20] Samuel P Costello, Patrick A Hughes, Oliver Waters, Robert V Bryant, Andrew D Vincent, Paul Blatchford, Rosa Katsikeros, Jesica Makanyanga, Melissa A Campaniello, Chris Mavrangelos, Carly P Rosewarne, Chelsea Bickley, Cian Peters, Mark N Schoeman, Michael A Conlon, Ian C Roberts-Thomson, and Jane M Andrews. Effect of Fecal Microbiota Transplantation on 8-Week Remission in Patients With Ulcerative Colitis: A Randomized Clinical Trial. JAMA, 321(2):156–164, January 2019. doi:10.1001/jama.2018.20046.

[21] Sudarshan Paramsothy, Ramesh Paramsothy, David T Rubin, Michael A Kamm, Nadeem O Kaakoush, Hazel M Mitchell, and Natalia Castaño-Rodríguez. Faecal Microbiota Transplantation for Inflammatory Bowel Disease: A Systematic Review and Meta-analysis. Journal of Crohn’s and Colitis, 11(10):1180–1199, October 2017. doi:10.1093/ecco-jcc/jjx063.

[22] Thomas J Borody and Annabel Clancy. Fecal microbiota transplantation for ulcerative colitis-where to from here? Translational gastroenterology and hepatology, 4, 2019. doi:10.21037/tgh.2019.06.04.

[23] David T Rubin, Ashwin N Ananthakrishnan, Corey A Siegel, Bryan G Sauer, and Millie D Long. ACG Clinical Guideline: Ulcerative Colitis in Adults. American Journal of Gastroenterology, March 2019. doi:10.14309/ajg.0000000000000152.

[24] Susannah J Salter, Michael J Cox, Elena M Turek, Szymon T Calus, William O Cookson, Miriam F Moffatt, Paul Turner, Julian Parkhill, Nicholas J Loman, and Alan W Walker. Reagent and laboratory contamination can critically impact sequence-based microbiome analyses. BMC Biology, 12(1):87, November 2014. doi:10.1186/s12915-014-0087-z.

[25] F Cold, P D Browne, S Günther, S I Halkjaer, A M Petersen, Z Al-Gibouri, L H Hansen, and A H Christensen. Multidonor FMT capsules improve symptoms and decrease fecal calprotectin in ulcerative colitis patients while treated - an open-label pilot study. Scandinavian journal of gastroenterology, 54 (3):289–296, March 2019. doi:10.1080/00365521.2019.1585939.

[26] Nathaniel D Chu, Jessica W Crothers, Le T T Nguyen, Sean M Kearney, Mark B Smith, Zain Kassam, Cheryl Collins, Ramnik Xavier, Peter L Moses, and Eric J Alm. Dynamic colonization of microbes and their functions after fecal microbiota transplantation for inflammatory bowel disease. bioRxiv, 2: 17004–36, January 2020. doi:10.1101/649384.

[27] Alka Goyal, Andrew Yeh, Brian R Bush, Brian A Firek, Leah M Siebold, Matthew Brian Rogers, Adam D Kufen, and Michael J Morowitz. Safety, Clinical Response, and Microbiome Findings Following Fecal Microbiota Transplant in Children With Inflammatory Bowel Disease. Inflammatory Bowel Diseases, 24(2):410–421, January 2018. doi:10.1093/ibd/izx035.

[28] David J Nusbaum, Fengzhu Sun, Jie Ren, Zifan Zhu, Natalie Ramsy, Nicholas Pervolarakis, Sachin Kunde, Whitney England, Bei Gao, Oliver Fiehn, Sonia Michail, and Katrine Whiteson. Gut microbial and metabolomic profiles after fecal microbiota transplantation in pediatric ulcerative colitis patients. FEMS Microbiology Ecology, 94(9):129, September 2018. doi:10.1093/femsec/fiy133.

[29] Mc Murdie, Paul J, Rosen Michael J, Han Andrew w, Johnson Amy Jo a, Holmes, Susan P, and Callahan Benjamin J. DADA2: High-resolution sample inference from Illumina amplicon data. Nature Methods, pages 1–7, May 2016. doi:10.1038/nmeth.3869.

[30] Ben J Callahan, Kris Sankaran, Julia A Fukuyama, Paul J McMurdie, and Susan P Holmes. Bioconductor Workflow for Microbiome Data Analysis: from raw reads to community analyses. F1000Research, 5, 2016. doi:10.12688/f1000research.8986.2.

[31] Kazutaka Katoh, George Asimenos, and Hiroyuki Toh. Multiple alignment of DNA sequences with MAFFT. Methods in molecular biology (Clifton, N.J.), 537(Suppl 5):39–64, 2009. doi:10.1007/978-1-59745-251-9_3.

[32] Price Morgan N, Dehal Paramvir S, and Arkin Adam P. FastTree 2–approximately maximum-likelihood trees for large alignments. PLoS ONE, 5 (3):e9490, March 2010. doi:10.1371/journal.pone.0009490.

[33] Lorenz Christian Reimer, Anna Vetcininova, Joaquim Sardà Carbasse, Carola Söhngen, Dorothea Gleim, Christian Ebeling, and Jörg Overmann. BacDive in 2019: bacterial phenotypic data for High-throughput biodiversity analysis. Nucleic acids research, 47(D1):D631–D636, January 2019. doi:10.1093/nar/gky879.

[34] Albert Barberán, Hildamarie Caceres Velazquez, Stuart Jones, Noah Fierer, and Steven J Hallam. Hiding in Plain Sight: Mining Bacterial Species Records for Phenotypic Trait Information. mSphere, 2(4): e00237–17, August 2017. doi:10.1128/mSphere.00237-17.

[35] Paul J McMurdie and Susan Holmes. phyloseq: an R package for reproducible interactive analysis and graphics of microbiome census data. PLoS ONE, 8(4):e61217. 2013. doi:10.1371/journal.pone.0061217.

[36] Adery C. A. Hope. A simplified Monte Carlo significance test procedure. Journal of the Royal Statistical Society, 30(3):582–598, 1968.

[37] Steven F Stoddard, Byron J Smith, Robert Hein, Benjamin R K Roller, and Thomas M Schmidt. rrnDB: improved tools for interpreting rRNA gene abundance in bacteria and archaea and a new foundation for future development. Nucleic acids research, 43(Database issue):D593–8, January 2015. doi:10.1093/nar/gku1201.

[38] The UniProt Consortium. UniProt: a worldwide hub of protein knowledge. Nucleic acids research, 47(D1):D506–D515, January 2019. doi:10.1093/nar/gky1049.

[39] Gavin M Douglas, Robert G Beiko, and Morgan G I Langille. Predicting the Functional Potential of the Microbiome from Marker Genes Using PICRUSt. Methods in molecular biology (Clifton, N.J.), 11(11): 169–177, 2018. doi:10.1007/978-1-4939-8728-3_11.

[40] Jessica D Forbes, Chih-Yu Chen, Natalie C Knox, Ruth-Ann Marrie, Hani El-Gabalawy, Teresa de Kievit, Michelle Alfa, Charles N Bernstein, and Gary Van Domselaar. A comparative study of the gut microbiota in immune-mediated inflammatory diseases-does a common dysbiosis exist? Microbiome, 6(1):221–15, December 2018. doi:10.1186/s40168-018-0603-4.

[41] Jessica Connors, Nick Dawe, and Johan Van Limbergen. The Role of Succinate in the Regulation of Intestinal Inflammation. Nutrients, 11(1):25, December 2018. doi:10.3390/nu11010025.

[42] G V Smirnova and O N Oktyabrsky. Glutathione in bacteria. Biochemistry. Biokhimiia, 70 (11):1199–1211, November 2005. doi:10.1007/s10541-005-0248-3.

[43] Susanna Nikolaus, Berenice Schulte, Natalie Al-Massad, Florian Thieme, Dominik M Schulte, Johannes Bethge, Ateequr Rehman, Florian Tran, Konrad Aden, Robert Häsler, Natalie Moll, Gregor Schütze, Markus J Schwarz, Georg H Waetzig, Philip Rosenstiel, Michael Krawczak, Silke Szymczak, and Stefan Schreiber. Increased Tryptophan Metabolism Is Associated With Activity of Inflammatory Bowel Diseases. Gastroenterology, 153(6):1504–1516.e2, December 2017. doi:10.1053/j.gastro.2017.08.028.

[44] Salvador Almagro-Moreno and E Fidelma Boyd. Insights into the evolution of sialic acid catabolism among bacteria. BMC evolutionary biology, 9(1): 118–16, May 2009. doi:10.1186/1471-2148-9-118.

[45] Mohammed A Alzoghaibi. Concepts of oxidative stress and antioxidant defense in Crohn’s disease. World journal of gastroenterology: WJG, 19(39): 6540–6547, October 2013. doi:10.3748/wjg.v19.i39.6540.

[46] Zhiqi Wang, Sai Li, Yu Cao, Xuefei Tian, Rong Zeng, Duan-Fang Liao, and Deliang Cao. Oxidative Stress and Carbonyl Lesions in Ulcerative Colitis and Associated Colorectal Cancer. Oxidative medicine and cellular longevity, 2016(3):9875298–15, 2016. doi:10.1155/2016/9875298.

[47] Jason Lloyd-Price, Cesar Arze, Ashwin N Ananthakrishnan, Melanie Schirmer, Julian Avila-Pacheco, Tiffany W Poon, Elizabeth Andrews, Nadim J Ajami, Kevin S Bonham, Colin J Brislawn, David Casero, Holly Courtney, Antonio González, Thomas G Graeber, A Brantley Hall, Kathleen Lake, Carol J Landers, Himel Mallick, Damian R Plichta, Mahadev Prasad, Gholamali Rahnavard, Jenny Sauk, Dmitry Shungin, Yoshiki Vázquez-Baeza, Richard A White, IBD-MDB Investigators, Jonathan Braun, Lee A Denson, Janet K Jansson, Rob Knight, Subra Kugathasan, Dermot P B McGovern, Joseph F Petrosino, Thaddeus S Stappenbeck, Harland S Winter, Clary B Clish, Eric A Franzosa, Hera Vlamakis, Ramnik J Xavier, and Curtis Huttenhower. Multi-omics of the gut microbial ecosystem in inflammatory bowel diseases. Nature, 569(7758):655–662, May 2019. doi:10.1038/s41586-019-1237-9.

[48] Tobias L Freitag, Anna Hartikainen, Hanne Jouhten, Cecilia Sahl, Seppo Meri, Veli-Jukka Anttila, Eero Mattila, Perttu Arkkila, Jonna Jalanka, and Reetta Satokari. Minor Effect of Antibiotic Pre-treatment on the Engraftment of Donor Microbiota in Fecal Transplantation in Mice. Frontiers in Microbiology, 10:2685, 2019. doi:10.3389/fmicb.2019.02685.

[49] K Ariake, T Ohkusa, T Sakurazawa, J Kumagai, Y Eishi, S Hoshi, and T Yajima. Roles of mucosal bacteria and succinic acid in colitis caused by dextran sulfate sodium in mice. Journal of medical and dental sciences, 47(4):233–241, December 2000.

[50] M Raithel, M Matek, H W Baenkler, W Jorde, and E G Hahn. Mucosal histamine content and histamine secretion in Crohn’s disease, ulcerative colitis and allergic enteropathy. International Archives of Allergy and Immunology, 108(2):127–133, October 1995. doi:10.1159/000237129.

[51] Elena García-Martin, Juan L Mendoza, Carmen Martínez, Carlos Taxonera, Elena Urcelay, José M Ladero, Emilio G de la Concha, Manuel Díaz-Rubio, and José A G Agúndez. Severity of ulcerative colitis is associated with a polymorphism at diamine oxidase gene but not at histamine N-methyltransferase gene. World journal of gastroen-terology: WJG, 4(4):615–620, January 2006. doi:10.3748/wjg.v12.i4.615.

[52] Shin-Hee Kim, Katharine G Field, Michael T Morrissey, Robert J Price, Cheng-I Wei, and Haejung AN. Source and Identification of Histamine-Producing Bacteria from Fresh and Temperature-Abused Albacore†. Journal of Food Protection, 64(7):1035–1044, July 2001. doi:10.4315/0362-028X-64.7.1035.

[53] Erica E Alexeev, Jordi M Lanis, Daniel J Kao, Eric L Campbell, Caleb J Kelly, Kayla D Battista, Mark E Gerich, Brittany R Jenkins, Seth T Walk, Douglas J Kominsky, and Sean P Colgan. Microbiota-Derived Indole Metabolites Promote Human and Murine Intestinal Homeostasis through Regulation of Interleukin-10 Receptor. The American journal of pathology, 188(5):1183–1194, May 2018. doi:10.1016/j.ajpath.2018.01.011.

[54] Teresa Zelante, Rossana G Iannitti, Cristina Cunha, Antonella De Luca, Gloria Giovannini, Giuseppe Pieraccini, Riccardo Zecchi, Carmen D’Angelo, Cristina Massi-Benedetti, Francesca Fallarino, Agostinho Carvalho, Paolo Puccetti, and Luigina Romani. Tryptophan catabolites from microbiota engage aryl hydrocarbon receptor and balance mucosal reactivity via interleukin-22. Immunity, 39(2):372–385, August 2013. doi:10.1016/j.immuni.2013.08.003.

[55] Yen-Lin Huang, Christophe Chassard, Martin Hausmann, Mark von Itzstein, and Thierry Hennet. Sialic acid catabolism drives intestinal inflammation and microbial dysbiosis in mice. Nature communications, 6:8141, August 2015. doi:10.1038/ncomms9141.

